# Expanding the phenotype spectrum of β-mannosidosis

**DOI:** 10.1101/2025.01.13.24316445

**Authors:** Angela Martin-Rios, Karolina M Stepien, Liliane H. Gibbs, Katherine Hall, Patricia L. Hall, Gisele Bentz Pino, Raymond Wang, Nishitha R. Pillai, Troy C. Lund, Paul J. Orchard, Virginia Kimonis

**Affiliations:** Division of Genetics, Department of Pediatrics, University of California - Irvine, Orange, CA, 92697, USA; Adult Inherited Metabolic Diseases Department, Salford Royal Organization, Northern Care Alliance NHS Foundation Trust, Salford, United Kingdom; Department of Radiological Sciences, School of Medicine, University of California, Irvine; CHOC Children’s Hospital, Orange, CA 92868, USA; Biochemical Genetics Laboratory, Division of Laboratory Genetics and Genomics, Mayo Clinic, Rochester, MN 55905, USA; Division of Metabolic Disorders CHOC Children’s Hospital, Orange, CA 92868; Department of Pediatrics, University of California-Irvine School of Medicine, Irvine, CA 92617, USA; Division of Genetics and Metabolism, Department of Pediatrics, University of Minnesota, Minneapolis, MN 55455, USA; Division of Blood and Marrow Transplantation, Department of Pediatrics, University of Minnesota, Minneapolis, MN 55455, USA; Department of Neurology, University of California - Irvine, Orange, CA, 92697, USA; Department of Pathology, University of California - Irvine, Orange, CA, 92697, USA

**Keywords:** β-mannosidase, *MANBA* gene, sensorineural hearing loss, intellectual disability, angiokeratoma corporis diffusum, abnormal brain white matter, hypomyelination

## Abstract

**Purpose:** To expand the spectrum of clinical and biochemical phenotypes, and brain imaging features of individuals with β-mannosidosis, a rare lysosomal storage disease.

**Methods:** We describe the clinical features of six patients with β-mannosidosis, their findings on brain magnetic resonance imaging (MRI), and the changes over time in two of them. We also review previously reported patients, analyze the variants in *MANBA,* the first symptom and spectrum of symptoms, and the ages of onset and of diagnosis of the disease.

**Results:** Forty-four patients have been reported to date, including our patients. The mean age of diagnosis is 12.8 years, and the age of onset of symptoms is 2.4 years. Hearing loss is the most frequently reported initial symptom and intellectual disability is the most frequent symptom overall. Erythromelalgia, nystagmus, macrocephaly, and obsessive-compulsive-like behavior are newly described features associated with β-mannosidosis. 40% of the patients have abnormal brain imaging. Brain MRI showed hypomyelination in one patient and abnormal white matter changes in another patient. Twenty-nine pathogenic variants in *MANBA* have been reported among the 44 patients; 60.6% of patients have private variants, and 39.4% have the recurrent variant c.2158-2A>G. A new oligosaccharide structure: Neu-Man2-GlcNac2, was found in the urine of two affected patients.

**Conclusion:** Due to disease heterogeneity, establishing a genotype-phenotype correlation remains challenging in β-mannosidosis. A spectrum of brain MRI abnormalities are described as manifestations of this condition: delayed myelination, hypomyelination, and abnormal white matter signals. Additional studies are needed to delineate the pathophysiology of this condition.

## INTRODUCTION

β-mannosidosis (OMIM #248510) is a lysosomal storage disorder caused by a deficiency of lysosomal β-mannosidase activity, which catalyzes the last step of glycoprotein degradation. The enzyme deficiency is caused by pathogenic variants in *MANBA* and has an autosomal recessive inheritance pattern.^1^ Along with α-mannosidosis (OMIM #248500), these diseases were initially referred to as oligosaccharidoses since they are suspected when oligosaccharides are detected in urine. α-mannosidosis is characterized by intellectual disability, frequent infections, immunodeficiency, hearing loss, coarse facies, and skeletal abnormalities, and has been widely described elsewhere.^2,3^ On the other hand, β-mannosidosis is very rare, and the exact incidence is unknown; however, the reported prevalence is 0.13 in the Netherlands^4^, 0.12 in Portugal ^5^, and 0.16 in the Czech Republic per 100,000 live births.^6^ This condition was first described in 1981 in Nubian goats,^7^ and later in Salers cattle^8^, dogs,^9,10^ and recently, in a domestic cat.^11^ In 1986, Cooper et al.^12^ and Wenger et al.^1^ described the first cases of β-mannosidosis in humans. Prior to this report, 38 patients have been described worldwide (Table 1). The clinical features and natural history of β-mannosidosis have been difficult to describe given the very low incidence of the disease and the heterogeneity of manifestations,^13^ including hearing loss,^14^ seizures,^15^ angiokeratomas,^16^ and different levels of developmental delay^17^ and intellectual disability.^18^ Availability of brain imaging in this condition is scarce, and there are no pathology reports of patients. Also, given the low number of patients reported, little information on the natural history of β-mannosidosis is available.

**Table 1.** Patients affected with β-mannosidosis reported in literature to date.

| n | Author and year | Sex | MANBA Variant 1 / Variant 2 (Reference of reported variant)<sup>†</sup> | Enzyme activity (source)<sup>‡</sup> |
| --- | --- | --- | --- | --- |
| 1 | Cooper, 1986<sup>12</sup> | M | c.247G>T, p.Glu83Ter / c.1276C>T, p.Gln426Ter (Bedilu, 2002)<sup>34</sup> | 0.8 (L), absent (P), 4 (F) |
| 2 |  | M | N/A | 0.2 (L) |
| 3 | Wenger, 1986<sup>1</sup> | M | N/A | Absent (L) |
| 4 | Dorland, 1988<sup>35</sup> | M | N/A | Absent (L, P) |
| 5 |  | M | N/A | Absent (L, P) |
| 6 | Kleijer, 1990<sup>18</sup> | F | c.2158-2A>G / c.2158-2A>G (Alkhayat, 1998)<sup>36</sup> | Absent (F) |
| 7 |  | M | c.2158-2A>G / c.2158-2A>G (Alkhayat, 1998)<sup>36</sup> | 0.9 (L) |
| 8 | Wijburg, 1992<sup>37</sup> | F | c.1454_1455del, p.Tyr485CysfsTer27 / c.1454_1455del, p.Tyr485CysfsTer27 (Bedilu, 2002)<sup>34</sup> | 4.7 (L), Absent (P, F) |
| 9 | Cooper, 1991<sup>38</sup> | F | N/A | 7.0 (L), 8.7 (F) |
| 10 | Poenaru, 1992<sup>39</sup> | M | c.375_378del, p.Tyr126AlafsTer11 / c.1513T>C, p.Ser505Pro (Riise Stensland, 2008)<sup>40</sup> | Absent (F), 0.5 (L), 0.1 (S) |
| 11 | Levade, 1994<sup>41</sup> | M | N/A | 3 (L), 13 (P) |
| 12 | Rodriguez-Serna, 1996<sup>42</sup> | F | c.1175G>A, p.Gly392Glu / c.1848del (Gort, 2006)<sup>31</sup> | 3 (L), 0.009 (S), 1.9 (F) |
| 13 | Gourrier, 1996<sup>43</sup> | F | N/A | Absent (L), 22 (S) |
| 14 | Uchino, 2002<sup>44</sup>, Suzuki, 2004<sup>45</sup> | M | c.960+1G>A / c.960+1G>A | 5.44 (P) |
| 15 | Cherian, 2004<sup>15</sup> | F | N/A | 33 (L), 18 (F), 21 (P) |
| 16 |  | F | N/A | 91 (L), 24 (F), 121 (P) |
| 17 | Sedel, 2006<sup>46</sup> | M | c.563_572dup, p.Trp192Ter / c.1705-1G>A | 1.08 (L), 1.4 (P) |
| 18 | Gort, 2006<sup>31</sup> | F | c.544C>T, p.Arg182Trp / c.1398G>A, p.Trp466Ter | Absent (L, S) |
| 19 | Molho-Pessach, 2007<sup>16</sup> | F | c.693G>A, p.Trp231Ter / c.693G>A, p.Trp231Ter | 0.2 (L), Absent (F), 1 (S) |
| 20 | Labauge, 2009<sup>47</sup> | M | c.1922G>A, p.Arg641His / c.1922G>A, p.Arg641His | 4 (L) |
| 21 | Broomfield, 2013<sup>48</sup> | F | c.293T>A, p.Leu98Ter / c.293T>A, p.Leu98Ter | 0.2 (L) |
| 22 | Blomqvist, 2019<sup>13</sup> | F | Chr4q24 13.1 kb inverted duplication (g.102669696_102669764ins102664572_102677720inv), homozygous | 0.06 (L) |
| 23 | Schrauen, 2019<sup>49</sup> | M | c.2158-2A>G / c.2158-2A>G |  |
| 24 | Safka Brozkova, 2020<sup>14</sup> | M | N/A | 1.65 (L) |
| 25-26-27 |  | M (2), F (1) | c.2158-2A>G / c.2158-2A>G |  |
| 28-29-30 |  | M (3) | c.2158-2A>G / c.2158-2A>G |  |
| 31 |  | F | c.2158-2A>G / c.2158-2A>G | Absent (L) |
| 32-33-34 |  | UNK (3) | c.2158-2A>G / c.2158-2A>G |  |
| 35 | Alshoraim, 2021<sup>50</sup> | M | N/A |  |
| 36 | Gowda, 2021<sup>17</sup> | M | c.1317+1G>A / c.1317+1G>A | Absent (S) |
| 37 |  | M | c.1317+1G>A / c.1317+1G>A | Absent (S) |
| 38 | He, 2022<sup>51</sup> | M | c.2538del, p.Met846IlefsTer27 / c.280C>A, p.Gln94Lys | 25.8 (L) |
| 39 | This publication | M | c.545G>A, p.Arg182Gln / c.545G>A, p.Arg182Gln | Absent (L) |
| 40 |  | M | c.575G>A, p.Trp192Ter / c.1499G>A, p.Arg500His | 2.62 (L) |
| 41 |  | F | c.1540_1541del, p.Val514CysfsTer10 / c.2352_2356del, p.Thr785LeufsTer27 | Absent (L) |
| 42 |  | F | c.1430A>G p.Tyr477Cys / c.1430A>G p.Tyr477Cys | Absent (L) |
| 43 |  | M | c.685C>T, p.Gln229Ter / c.685C>T, p.Gln229Ter | 3 (S) |
| 44 |  | F | c.1452_1453del, p.Tyr485CysfsTer27 / c.1753C>T, p.Arg585Ter | 2 (P), Absent (L) |

\* Number in parenthesis are the reported patients in the correspondent publication. UNK: Unknown.
<sup>†</sup> RefSeq NM\_005908.4, GRCh38. N/A= not available.
<sup>‡</sup> For β-mannosidase enzyme activity in leukocytes (L) and fibroblasts (F), the units are in nmol/h/mg. For serum (S) and plasma (P), the units are in nmol/h/ml.

Here, we describe six patients with a confirmed diagnosis of β-mannosidosis, with varying clinical manifestations and severity. We also describe the brain magnetic resonance imaging (MRI) of all six patients, three of whom have white matter changes. Brain imaging findings have been reported previously in one individual^19^ included in our present cohort (Patient 2). We also describe the evolution of β-mannosidosis in this patient, the brain MRI changes over time, and his current condition. Finally, we review the previously reported patients, their clinical features, and the associated *MANBA* variants.

## MATERIALS AND METHODS

### Assessment of MANBA variants and effects on the protein

The *MANBA* variants of each patient recruited were initially analyzed using Mutation Taster ^20^ and Polyphen 2 ^21^ for pathogenicity prediction. We analyzed the missense variants of the patients with Clustal Omega^22^ for protein alignment. The protein sequences used for alignment were: NP_005899.3 (*Homo sapiens*), XP_001169005.2 (*Pan troglodytes*), XP_028704480.1 (*Macaca mulatta*), NP_081564.3 (*Mus musculus*), NP_776812.1 (*Bos taurus*), XP_005639293.1 (*Canis lupus familiaris*), NP_001272620.1 (*Capra hircus*), XP_040527136.1 (*Gallus gallus*), NP_956453.1 (*Danio rerio*), NP_510342.1 (*Caenorhabditis elegans*). The variants were located on the schematic diagram built with IBS 2.0 ^23^, and the 3D reconstructions were done using Alphafold ^24^ and UCSF Chimera X (https://www.cgl.ucsf.edu/chimerax/index.html) ^25^ based on the published databases (Uniprot, entry access O00462 Beta mannosidase) ^26^.

### Brain MRI analysis

Magnetic resonance images (MRI) from the brain obtained for each patient were reviewed by a pediatric radiologist (LG) with expertise in pediatric neurologic conditions. Changes over time were also analyzed for the available images.

### Urine oligosaccharide analysis

Urine oligosaccharide profiles were performed for patients 1, 2, 3 and 4 by matrix assisted laser desorption ionization – time of flight (MALDI-TOF) mass spectrometry, as described by Xia et al.^27^

### Review of literature

Original research and case report papers were included in the review of literature, and a list of reported patients was obtained (Table 1). The terms used for Pubmed search were “beta mannosidosis,” “MANBA,” “beta mannosidase,” “human,” “patient,” and “case report.” The reported variants of each patient were updated to the GRCh38 (RefSeq NM_005908.4) and validated accordingly.^28^ For each reported patient, a thorough review of listed symptoms was made and classified into present (+), absent (-), and not reported (NR) (Table S1). The symptoms included in the study were based on the most frequent symptoms associated with β-mannosidosis reported in IEMbase,^29^ including developmental delay, intellectual disability, hearing loss, angiokeratoma, facial dysmorphism, skeletal dysplasia, behavioral abnormalities, organomegaly, ataxia, and seizures. Recurrent infections were included as a previously reported symptom associated with β-mannosidosis.^30^ Descriptive statistics, including mean, standard deviation, range, frequencies, and correlation, were made using SPSS version 29.0.0.0. The reported variants were also displayed schematically using IBS 2.0 (Fig. 1)^23^.

**Figure 1.**
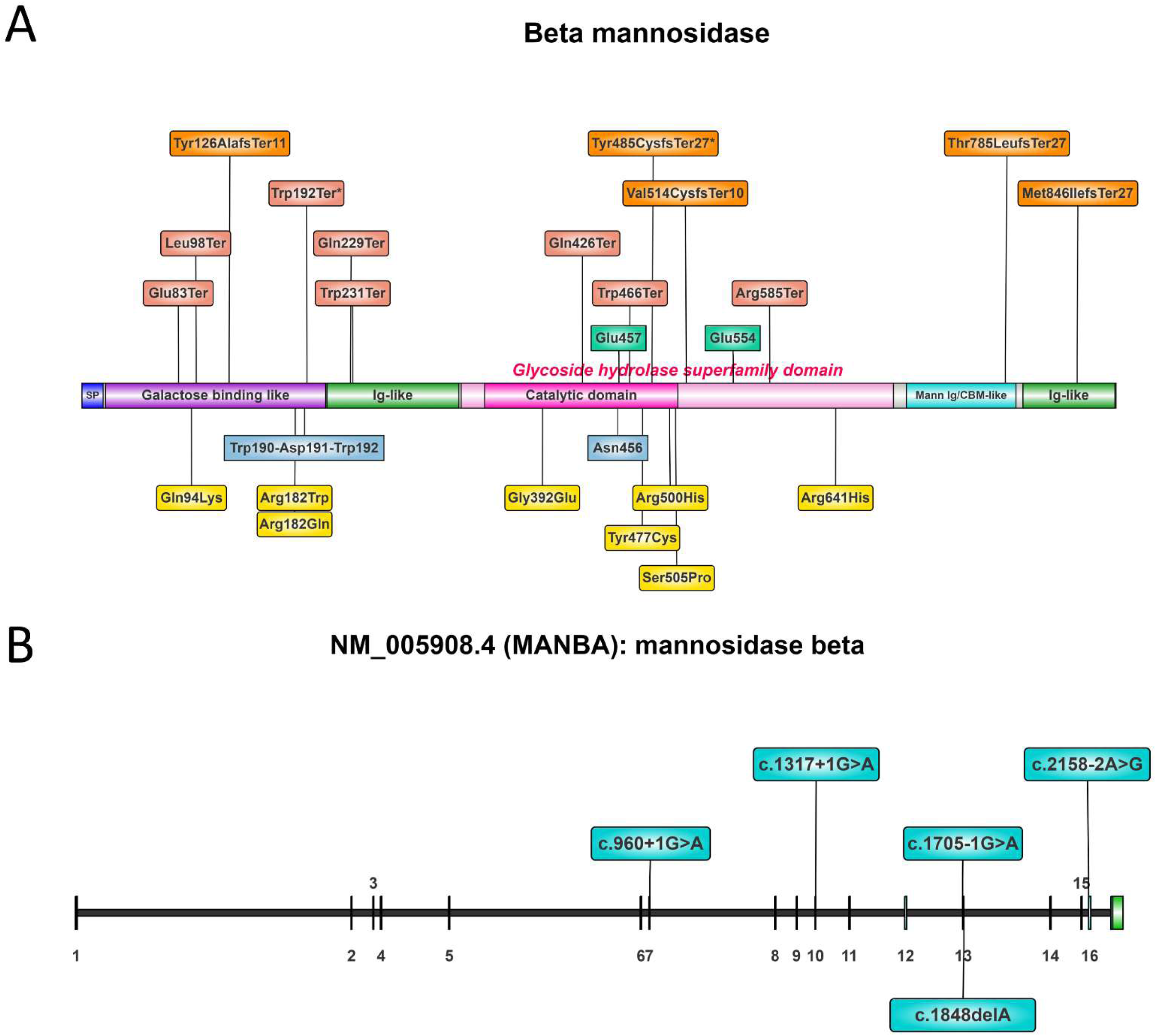
**A.** Schematic illustration of beta mannosidase protein domains and location of the reported mutations: frameshift mutations in orange, nonsense mutations in red, missense mutations in yellow. The residues in the aqua boxes are the active sites, and the blue boxes are the binding sites of the protein. **B.** Schematic illustration of *MANBA* gene, and the splice site mutations reported in literature.

## DESCRIPTION OF THE PATIENTS

### Patient 1

Patient 1 is a male with bilateral sensorineural hearing loss detected before 5 years old. Pregnancy was uneventful, and he was born by an emergency cesarean section at 38 weeks due to heartbeat decelerations. Birth weight and head circumference were normal; length was 45.7 cm (1^st^ centile). Newborn screening was normal. Motor milestones were normal; language was delayed. There is no history of metabolic decompensation or neurological regression. He has learning difficulties associated with anxiety, obsessive-compulsive traits, and perseveration. The patient is also being evaluated for precocious puberty under 5 years of age. Family history was negative for deafness or intellectual disability. Although both parents belonged to the same geographic region, consanguinity was denied. During physical examination, the height was 132 cm (83^rd^ centile), weight was 43.1 kg (99^th^ centile), and there were no dysmorphic features, angiokeratoma, or skeletal malformations. There was no gynecomastia, axillary hair or odors, or liver or spleen enlargement. Tanner stage was 1. The left testicle was 1-2 ml, and the right testicle was 3 ml in volume.

A hearing loss genetic panel showed a homozygous missense variant in *MANBA* (NM_005908.4:c.545G>A, p.Arg182Gln, Fig.S1A) initially classified as a variant of uncertain significance (VUS). Oligosaccharides in urine showed excretion of mannosyl-β(1 → 4)-N-acetylglucosamine and sialyl-α(2 → 6)-mannosyl-β(1 → 4)-N-acetylglucosamine, typical of β-mannosidosis (Fig. 2). The enzyme activity of β-mannosidase in leukocytes was completely absent. *In silico* prediction classifies this variant as disease-causing (Mutation Taster) and probably damaging (Polyphen 2, score 1.0). Arg182 is a highly conserved residue; and the 3D structure of this variant shows that Arg182 has six hydrogen bonds with Met122, Met416, and Glu435 residues; however, Gln182 only has one hydrogen bond with Met122. The residues Met416 and Glu435 are located on the catalytic domain of MANBA (Fig.S1A). An affected patient with β-mannosidosis was previously reported with the variant Arg182Trp, the same codon of this patient ^31^. This variant disrupts the interface between the galactose binding-like domain and the glycoside hydrolase superfamily domain ^32^, and this effect is seen in the Arg182Gln variant. In this context, with an absent enzyme activity and elevated excretion of oligosaccharides, the patient was diagnosed with β-mannosidosis, and the variant was reclassified.

**Figure 2.**
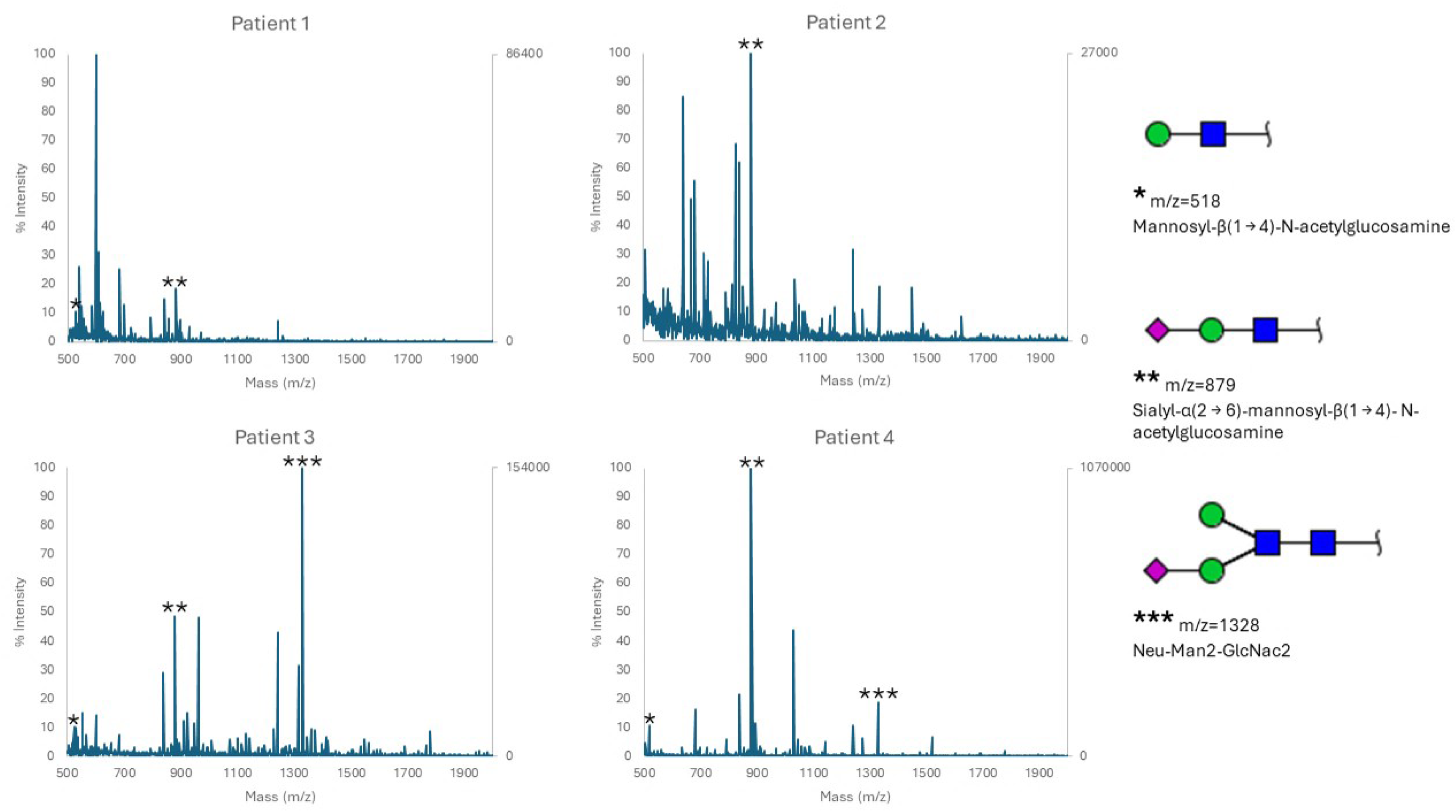
Urine oligosaccharide profiles of Patients 1 to 4 with β-mannosidosis. Characteristic peaks are labeled, and theoretical structures are provided in the inset. Patient 2 is post-transplant and only one characteristic peak (m/z=879) is present in the profile. Patients 3 and 4 have a third peak (m/z=1328), corresponding to a Neu-Man2-GlcNac2 structure.

### Patient 2

This male patient was previously reported by Lund et al, 2019.^33^ Pregnancy was overall uneventful, however, due to preeclampsia, the mother was induced, and delivery was at 38 weeks via c-section due to failure to progress. The motor milestones were noted to be delayed as well as speech. Macrocephaly was detected between 0-1 year and a brain MRI showed mild to moderate cerebral delayed myelination. The patient was diagnosed with autism spectrum disorder, and metabolic tests (ammonia, lactic acid, pyruvic acid, plasma amino acids, plasma acylcarnitine profile, urine amino acid profile, and urine organic acid profile) were reported as normal. Fragile X trinucleotide repeats and chromosomal microarray were also normal per report. Between 1 and 5 years of age the patient displayed progressive loss of muscle strength. An enzyme activity panel for lysosomal storage disorders showed low β-mannosidase activity (2.62 nmol h^-1^ mg^-1^, normal range 10–162.4 nmol h^-1^ mg^-1^). Sequencing of *MANBA* showed one likely pathogenic variant (c.575G>A, p.Trp192Ter) and a VUS (c.1499G>A, p.Arg500His) that was predicted as deleterious by Mutation Taster and Polyphen2. The Arg500 residue is highly conserved, and although Arg and His are positive charged amino acids, the His500 variant destabilizes the catalytic domain of the protein because it has an imidazole group that creates an abnormal hydrogen bond with Ser499, causing the loss of hydrogen bonds that Arg originally has with Ile449, His446 and Lys444 (Fig.S1B).

The patient received a HLA matched umbilical cord blood transplant (a detailed description is available in Lund et al.^33^). At puberty, his weight was 44.5 kg (29^th^ centile), height was 172.5 cm (91^st^ centile), and BMI 14.94 (1^st^ centile). Physical examination showed mild coarse and dysmorphic features (thick lips, bilateral epicanthal folds), bilateral elliptical nystagmus, slender fingers, and scoliosis. There was no hepatosplenomegaly or angiokeratomas. Gait was ataxic requiring a walker, ankle foot orthosis braces or a wheelchair for mobilization. After the transplant, the levels of β-mannosidase enzyme activity in leukocytes were normalized; near normal levels in dried blood spots and plasma were confirmed, and sialyl-α(2 → 6)-mannosyl-β(1 → 4)-N-acetylglucosamine was the only oligosaccharide found in urine (Fig.2).

### Patient 3

The patient is a female with an uneventful prenatal and birth history and normal hearing and metabolic newborn screening. Between 1 and 5 years of age the patient had multiple upper respiratory infections that required nebulizer and/or inhalators, requiring adenoidectomy and tonsillectomy. At the same age, the patient was noted to have delayed language for her age, and an audiology evaluation led to the detection of bilateral sensorineural hearing loss, resulting in the use of hearing aids. Her motor milestones were normal. Between 1-5 years of age, she developed behavior issues and frequent tantrums, and later the patient developed seizures, consisting in deviation of eyes to the left, mild eye twitching, drooling, and lack of verbal response to stimuli. Strabismus and bilateral optic nerve damage were also diagnosed at this age. Due to the findings on brain MRI suggestive of metabolic white matter disease, a lysosomal activity panel was requested, which showed absent β-mannosidase activity. Sequencing of *MANBA* gene showed two frameshift variants, c.1540_1541delGT, p.Val514CysfsTer10 maternally inherited, and c.2352_2356delGACCA, p.Thr785LeufsTer27 paternally inherited (Fig. 1). At puberty, the patient developed a change in the semiology of her seizures, consisting of multiple and recurrent episodes of tonic-clonic movements lasting 1-2 minutes, with 5 minutes of resolution in between, which required medical attention in the emergency room for stabilization, and posterior adjustment of medication for seizure control. She returned to her baseline health status but developed urinary incontinence after the episode.

At the last evaluation, the patient was unable to perform her daily activities independently and developed aggressive behavior. Weight was 63.7 kg (86^th^ centile), height was 162.6 cm (59^th^ centile), and BMI was 27.47 (87^th^ centile). The physical examination showed a girl with mild dysmorphic features: esotropia, broad nose bridge, flat philtrum, microretrognathia, and mildly prominent cheeks. The hands revealed slender fingers with bilateral clinodactyly of the fifth finger. Her palmar creases were shallow, and there was a single palmar crease of the right hand. The skin was fair, and she did not have angiokeratomas. There was no skeletal malformations or hepatosplenomegaly. The urine oligosaccharides showed excretion of mannosyl-β(1 → 4)-N-acetylglucosamine and sialyl-α(2 → 6)-mannosyl-β(1 → 4)-N-acetylglucosamine. A third oligosaccharide was found, corresponding to a Neu-Man2-GlcNac2 structure (fig. 2).

### Patient 4

This patient is a female born at 34 3/7 weeks via c-section due to severe intrauterine growth restriction. Pregnancy was complicated by maternal use of sertraline and possible maternal use of tetrahydrocannabinol. Birth weight was 1490 g (<1^st^ centile), length was 37 cm (<1^st^ centile), head circumference was 28 cm (<1^st^ centile), and APGAR scores of 1 and 5 at one and five minutes, respectively. She required continuous positive airway pressure (CPAP) support due to poor respiratory effort, and phototherapy due to jaundice. She was hospitalized in the NICU for one month. During physical examination, bifid uvula was found in addition to a parasacral dimple with asymmetrical gluteal cleft. Submucous cleft palate and lumbar spinal cord malformation were suspected. A cleft palate and laryngeal cleft were confirmed and repaired. A lumbar spine MRI showed a low-lying spinal cord with tip at L3, and a sinus tract that runs inferiorly to end at coccyx.

A chromosomal microarray revealed no copy number variants but regions of homozygosity approximating 26% of her total DNA.

During the first year the patient had ear infection and recurrent diarrhea, and a BAER and audiometry confirmed bilateral moderate to severe sensorineural hearing loss, for which hearing aids were placed. Developmental delay was also detected, with lack of speech but ability to learn sign language. She developed staring spells, in which she had no response to voice or tactile stimulation. An electroencephalogram was normal.

Exome sequencing performed at this time detected two sets of homozygous variants in the *MANBA* gene. The first variant, c.434A>C, p.Gln145Pro, was classified as VUS. Mutation Taster classifies this variant as a polymorphism, and Polyphen2 classifies it as possibly damaging (Score 0.611). The Gln145 residue is conserved only in mammals, except for mouse (Fig.S1C). This residue creates two hydrogen bonds with residues, while Pro does not make any bonds. However, the residues in contact with Gln145 belong to the same domain, therefore, it is inferred that this change will not cause a major disruption on the tridimensional structure of the protein. The second variant, c.1430A>G, p.Tyr477Cys, was classified as VUS. Mutation Taster and Polyphen2 classified this variant as damaging. It was reclassified as likely pathogenic. The Tyr477 residue is highly conserved, and the Cys477 change disrupts the contact between the galactose-binding-like domain and the catalytic domain, creating an abnormal hydrogen bond with Asp473, and eliminating the bond originally created between Tyr477 and Gln162 (Fig.S1D). The enzyme activity level of β-mannosidase in leukocytes was absent, confirming the diagnosis of β-mannosidosis. The urine oligosaccharides profile showed three abnormal peaks, corresponding to excretion of mannosyl-β(1 → 4)-N-acetylglucosamine, sialyl-α(2 → 6)-mannosyl-β(1 → 4)-N-acetylglucosamine, and a Neu-Man2-GlcNac2 oligosaccharide structure (fig.2).

Between 1 and 5 years of age the patient developed episodes of recurrent pain in hands and feet, and bright red-hot swelling in the setting of activity, consistent with erythromelalgia. Electromyography and nerve conduction tests were normal. At this age a gastrostomy was placed due to chronic diarrhea and poor oral intake, and she was diagnosed with autism spectrum disorder. The patient developed progressive dysphagia, with inability to swallow solids or liquids, requiring nutrition exclusively through gastrostomy. The family history is relevant for a high degree of consanguinity. At physical examination, her weight was 12.8 kg (38^th^ centile), height was 86.5 cm (10^th^ centile), and BMI was 17.11 (80^th^ centile). Dysmorphology evaluation was significant for mild coarseness of the face, long palpebral fissures, bilateral epicanthal folds, and flat nose bridge. Ears were normal. Uvula was broad, and palate was intact. The skin and hair were fair, very lightly pigmented, and there were angiokeratomas of all her limbs. There was no hepatosplenomegaly and no skeletal deformities or dysplasia. There was axial hypotonia, with normal strength and reflexes in limbs.

### Patient 5

The patient is a male in his twenties born to healthy parents following a normal pregnancy and delivery. Birth weight was 2.75 kg at 37 weeks’ gestation. The developmental milestones were reached at a normal time, with no history of neuroregression. Hearing loss was detected between 1-5 years old, and learning difficulties detected between 6-10 years old. The symptoms have remained stable throughout his adult life. He is being treated with methylphenidate due to behavioral issues and attention deficit and hyperactivity disorder (ADHD) symptoms, and he has had panic attacks. There is no history of seizures, balance problems or ataxia. He also is being followed for primary hypogonadism. The patient was diagnosed with β-mannosidosis at puberty, and his two siblings have the same condition.

At the last review at adulthood, his weight was 39.5 kg (4^th^ centile), height was 158 cm (4^th^ centile), and BMI was 16 kg/m2. The patient had subtle coarse facial features. He had angiokeratomas of the thighs, buttocks, groin, palms, and soles. The abdomen revealed both liver and spleen palpable at 2 cm. There was no tremor in his upper limbs and reflexes were normal. There were no visible skeletal malformations.

Urine oligosaccharides chromatography showed a band running in line with the disaccharides control band. The plasma β-mannosidase enzyme activity was 1 nmol/ml/h (normal range 150-1500), and leukocyte β-mannosidase activity was 0 nmol/ml/hr. *MANBA* sequencing detected a homozygous c.685C>T, p.Gln229Ter variant (Fig.1), later reclassified as pathogenic. This variant has not been reported in the published literature and has not been detected in control individuals from the gnomAD dataset2.

### Patient 6

The patient is a female in her thirties with hearing problems between 1-5 years old and requiring hearing aids between 6-10 years old. She was born at 36 weeks and her birth weight was 4 pounds 8 ounces (2177 g, <1^st^ centile). In addition to developmental delay, she has been diagnosed with intellectual disability. Assessment of her cognitive ability using British Abilities Scales II suggested scores in the low range; verbal score 68SS-2^nd^ centile; non-verbal reasoning score 85 SS-16^th^ centile; special score 62 SS <1^st^ centile; general conceptual ability 64 to 76-2^nd^ centile. The patient also had a diagnosis of ventricular septal defect that required surgery before one year of age, and she takes irbesartan 75 mg daily. Her significant medical problems include anxiety and disturbed sleep, which affects the family, some autistic features, and frequent tremor in upper limbs. There was no history of behavioral problems, aggressivity or seizures. She has chronic anemia due to abnormal uterine bleeding.

The physical examination showed a female patient with no obvious dysmorphic features and no skeletal abnormalities. Mild angiokeratomas were found on her palms. Height was 160 cm and weight was 67 kg. There was no tremor in upper limbs. A loud murmur in her aortic area and her chest was heard; blood pressure was 141/94. There was no hepatosplenomegaly. The evaluation by physical therapy reported independent walking with unorthodox wide arm swing, ataxia and hyperlordotic posture without scoliosis.

Exome sequencing revealed two variants in *MANBA*: c.1452_1453del, p.Tyr485CysfsTer27 and c.1753C>T, p.Arg585Ter (Fig.1). The oligosaccharides excretion in urine showed a disaccharide band present consistent with the diagnosis of β-mannosidosis. The plasma β-mannosidase enzyme activity was 2 nmol/ml/h (150-1500) and 0 nmol/mg/h in leukocytes.

## BRAIN MRI FINDINGS

***Patient 1 (Fig. S2A).*** MRI reports completely normal brain imaging. The spectroscopy was normal (not shown).

***Patient 2 (Fig. 3).*** This patient was followed with brain MRI every two months after the bone marrow transplant. The first imaging available before transplant shows T2 hyperintensities corresponding to abnormal myelination located at the periventricular and subcortical white matter in a symmetric fashion, including the posterior limb of the internal capsule, and a posteriorly deformed corpus callosum. The cerebellum, diploe and sinuses do not show abnormalities. The SWI sequence was normal. Figure 3A shows the sequential axial projections in FLAIR over time between 1 and 15 years of age. The most recent MRI demonstrates mild cortical atrophy along with the slow progression of the white matter changes, which are more extensive and symmetrical in the frontal and occipital areas. There is progressive thinning of the corpus callosum, and there are no changes in the cerebellum or brainstem.

**Figure 3.**
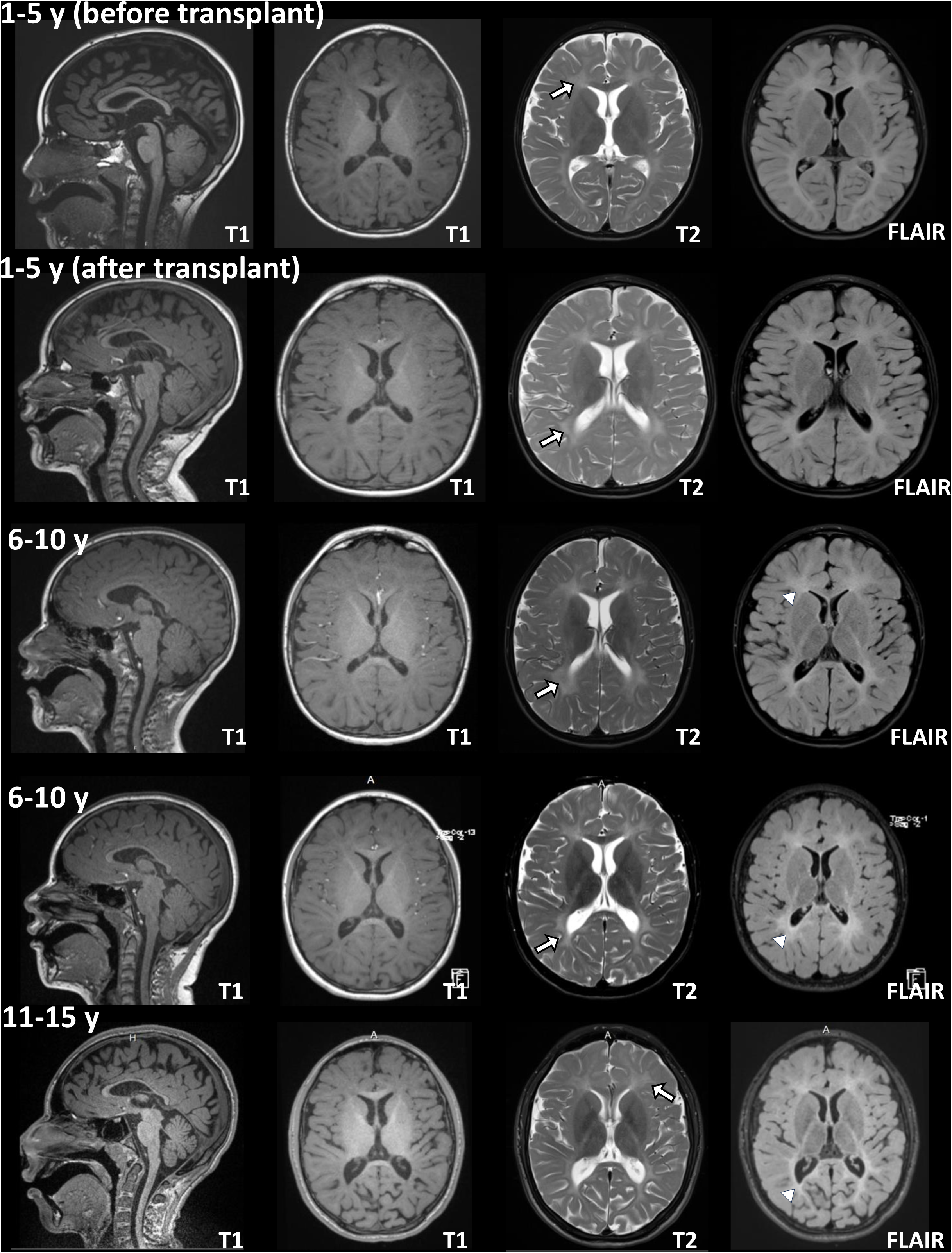
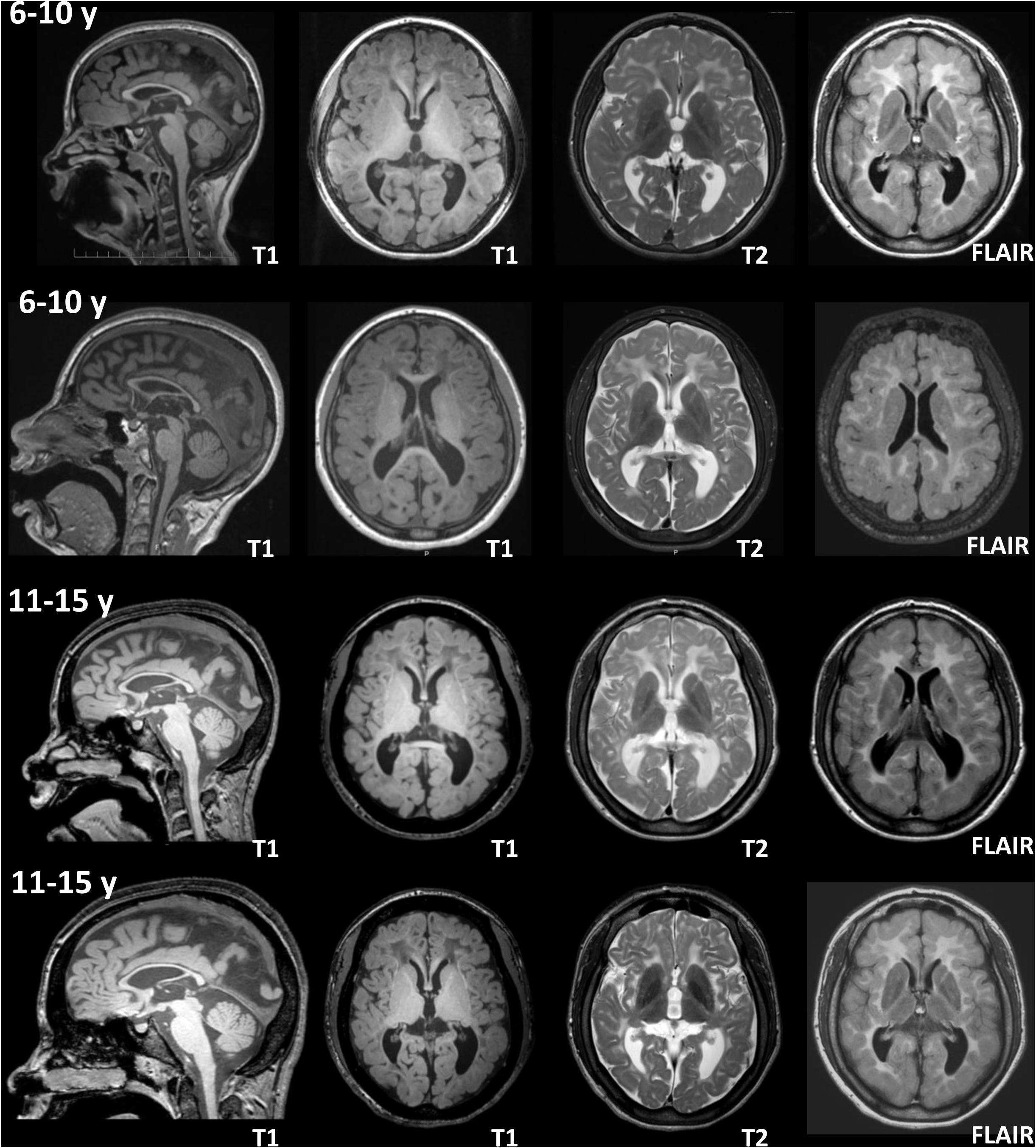
MRI imaging of Patient 2. Serial brain MRIs before transplant and between 5 and 15 years of age. From left to right for each row of images the projections are sagittal T1, axial T1, axial T2 and axial FLAIR. Signs of hypomyelination are evident with mild T2 hyperintensity in the periventricular white matter noted particularly on T2 axial images (arrows) and FLAIR sequences (arrowheads), with no significant change overtime. Mild posterior corpal callosal atrophy is also noted and unchanged overtime.

***Patient 3 (Fig.4).*** Significant findings in the first brain MRI included markedly abnormal white matter, indicated by a diffuse increased T2 signal involving both hemispheres symmetrically, predominantly in the frontal lobe and subcortical areas, without compromise of the anterior and posterior limbs of the internal capsule. The left caudate body exhibits some lobularity with increased signal intensity. Cortical atrophy is evident, with prominence of the sulci and ventricles. The cerebellum and brainstem are unaffected. Brain MRIs between 5 and 15 years of age did not show major changes compared to the first MRI except for thinning of the corpus callosum. MRI spectroscopy was normal in the left basal ganglia and the left frontal white matter.

**Figure 4.**
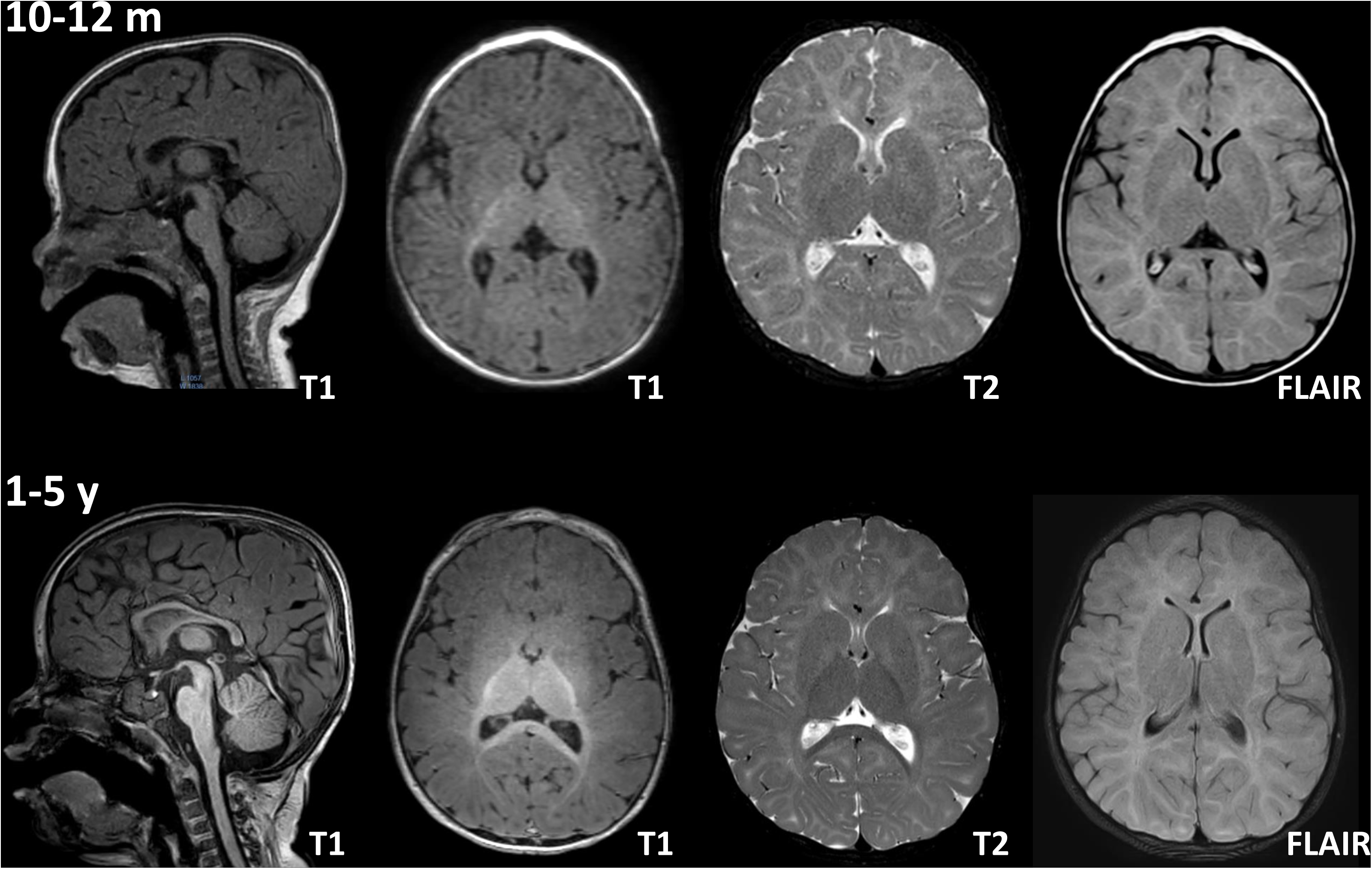
Brain MRI of Patient 3 at four time points between 5 and 15 years of age. From left to right for each row of images the projections are sagittal T1, axial T1, axial T2 and axial FLAIR. There is diffuse and symmetric abnormal increase in white matter T2 signal that extends from the deep periventricular white matter to the subcortical region. There are also signs of cerebral volume loss given by the prominence of the sulci and ventricles without substantial change overtime. The corpus callosum has normal signal and shape between 5-10 years old, and progressive thinning is evident between 11 and 15 years old.

***Patient 4 (Fig. 5).*** The first brain MRI of this patient before the 1^st^ year of age revealed severely delayed myelination for age; however, it was possible to see incipient myelination on T1 on the brain parenchyma. The second MRI between 1 and 5 years old reveals a progression of myelination in the cerebral white matter compared to the previous MRI, albeit delayed for age. The myelination is greater in parietal and occipital regions, with relative paucity of margination of the anterior limbs of the internal capsules and the frontotemporal white matter.

**Figure 5.**
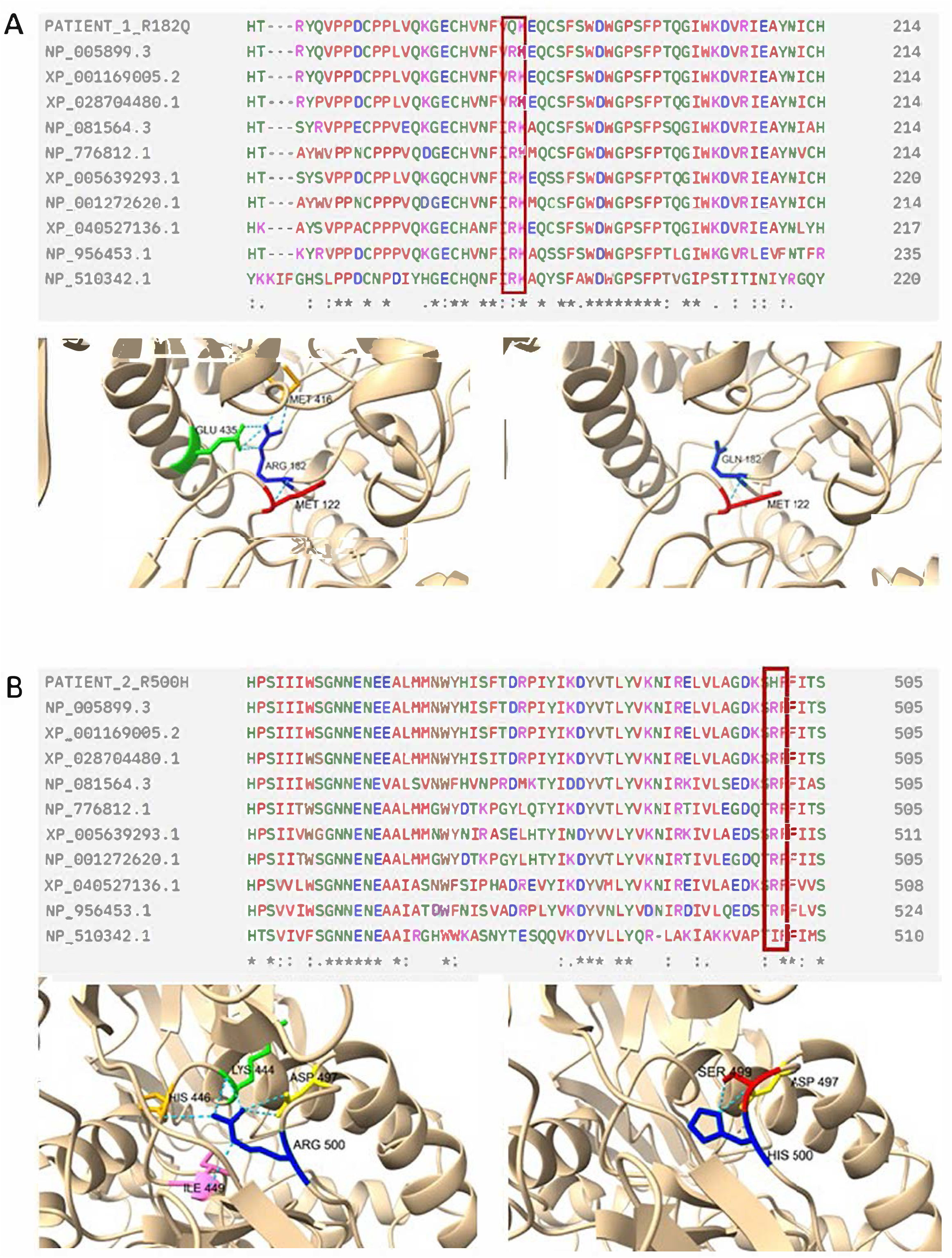
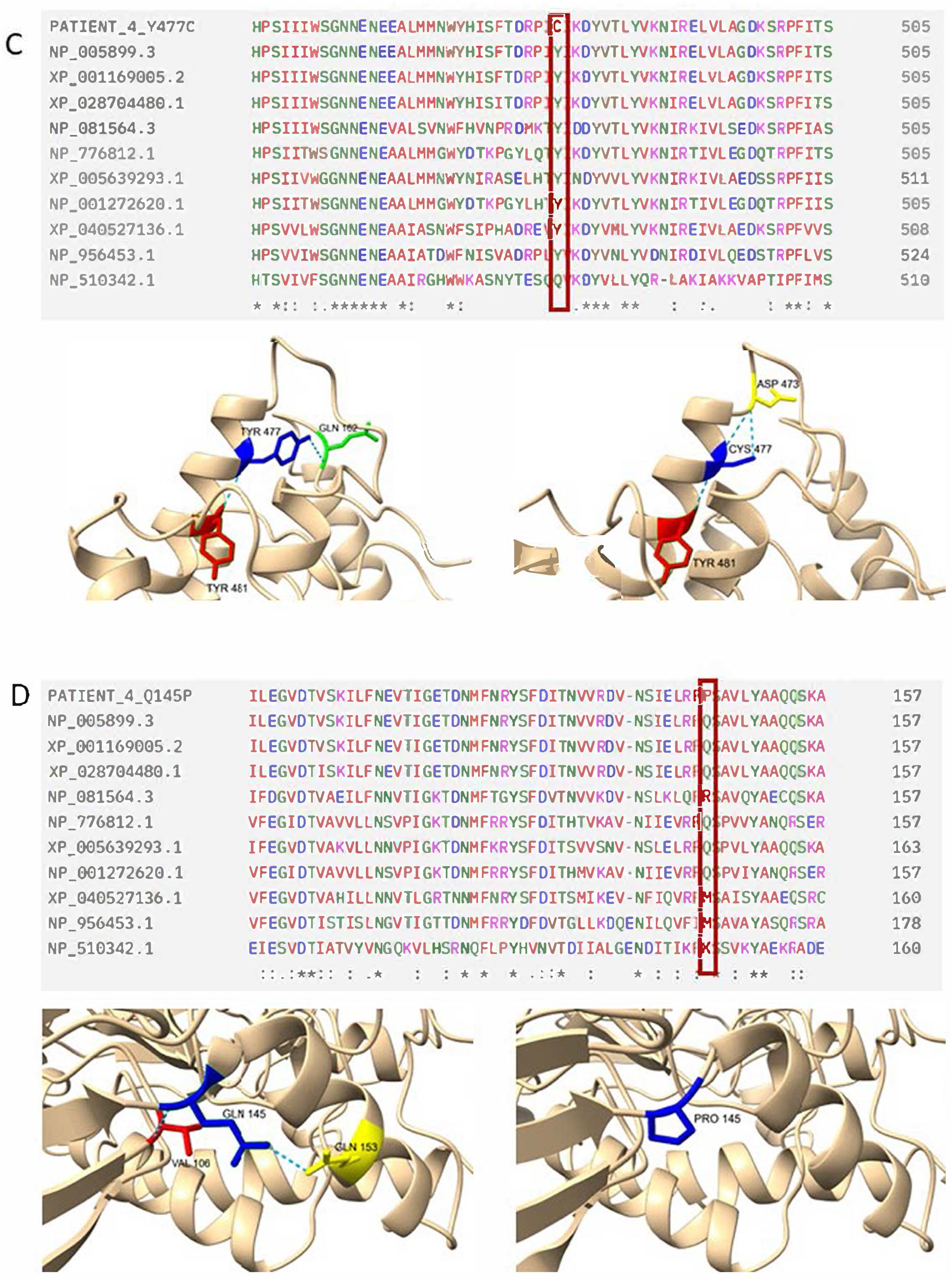
Brain MRI of Patient 4 before 1^st^ year and between 1-3 years of age. From left to right for each row of images the projections are sagittal T1, axial T1, axial T2 and axial FLAIR. The MRI obtained under 12 months of age, shows minimal myelination of the posterior limbs of the internal capsule (Axial T1) without substantial myelin in the lateral thalami or in the central corona radiata which are usually seen even at birth. Although the myelination has slightly progressed, it is still delayed for age.

***Patient 5 (Fig. S2B).*** The brain MRI of this patient at showed normal imaging findings.

***Patient 6 (Fig. S2C)*** The brain MRI is normal.

## REVIEW OF LITERATURE

Table 1 shows a list of the patients with β-mannosidosis reported in literature. For the analysis, we included the six patients reported in this paper, and only the data specifically reported in each scientific paper were included in the analysis.

To date of this publication, 44 patients have been described in the literature: 17 females (41.5%), 24 males (58.5%), and three patients in which sex was not reported. The mean age of diagnosis was 12.87 years (range 5 months – 51 years). The mean age of onset of symptoms was 2.39 years (28.7 months, range 0-12 years). The average time between the onset of symptoms and the diagnosis for the available data was 10.3 years (range 3 months – 49 years). In 32 patients, hearing loss (8 patients, 25%) was the most frequent first symptom reported (Table 2, table S1). Interestingly, angiokeratoma corporis diffusum was found as a first symptom in only two patients (6.3%) although it is frequently reported in β-mannosidosis (52.6%, table 2). We also evaluated the most frequently reported symptoms independently of age of onset (Table 2, table S1). The most frequent symptom was intellectual disability (91.4%), followed by behavioral abnormalities (82.1%) and hearing loss (74.4%). Compared to α-mannosidosis in which skeletal dysplasia is a main feature,^3^ in this review it was found only in 7.4% of the patients with β-mannosidosis.

**Table 2.** Frequency of the first symptom and most frequent symptom associated with β-mannosidosis reported in literature.

| First symptom | n (%) | Most frequent symptom | n (%) |
| --- | --- | --- | --- |
| Hearing loss | 8 (25) | Intellectual disability | 32 (91.4) |
| Developmental delay | 6 (18.8) | Behavioral abnormalities | 23 (82.1) |
| Intellectual disability | 4 (12.5) | Hearing loss | 29 (74.4) |
| Feeding difficulties | 3 (9.4) | Recurrent infections | 17 (70.8) |
| Seizures | 3 (9.4) | Developmental delay | 18 (56.3) |
| Speech delay | 3 (9.4) | Angiokeratoma | 10 (52.6) |
| Angiokeratoma | 2 (6.3) | Facial dysmorphism | 13 (41.9) |
| Frequent infections | 2 (6.3) | Seizures | 6 (40) |
| Apathy, disinterest. Hand weakness. | 1 (3.1) | Organomegaly | 4 (16) |
| Total | 32 (72.7%) | Ataxia | 4 (13.8) |
| Not reported | 12 (27.3%) | Skeletal dysplasia | 2 (7.4) |

The brain findings on computed tomography (CT) and magnetic resonance (MRI) were described in 39.4% of the patients (15/38 patients, not including the patients of our cohort), and it was normal in 9 (60%) and abnormal in 6 (40%). The described findings on CT included cerebral atrophy,^38^ basal ganglia and white matter calcification,^45^ and hydrocephalus.^47^ On brain MRI the findings included cortical and subcortical atrophy,^47^ periventricular and subcortical white matter hyperintensities,^17^ hydrocephalus^48^ and delayed myelination.^13^

There are 29 different pathogenic variants in *MANBA* associated with β-mannosidosis reported in literature thus far (Table 1). From them, 7 variants (24.1%) are frameshift (6 deletions, 1 insertion), 8 variants (27.5%) are missense, 8 variants (27.5%) are nonsense, 5 variants (13.7%) are in splice sites, and one (3.4%) is a chromosome inverted duplication encompassing 13.1 kb. The majority of the patients with reported variants have private mutations (20/33, 60.6%), and the patients that share variants belong to the same family.^17^ Only 13 patients (39.4%), all with Roma ancestry (12 from Czech Republic/Slovakia, 1 from Hungary) share a common variant (c.2158-2A>G) in homozygous status.

Figure 1 shows the location of the variants in the protein (A) and the splicing variants in the gene (B). Seven variants are located in the galactose-binding-like domain, and eight in the catalytic domain. There are three positions with recurrent variants: Arg182, Trp192 and Tyr485. The codon Arg182 has two missense variants reported (Arg182Trp^31^ and Arg182Gln, Patient 1). The codon Trp192 belongs to the binding site of *MANBA*, and has one nonsense variant (c.575G>A, p.Trp192Ter, Patient 2) and one frameshift variant (c.562_571dup, Trp192Ter).^46^ The codon Tyr485 is located in the catalytic domain of the protein, and has two frameshift variants (c.1454_1455del,^34^ and c.1452_1453del, Patient 6) with the same effect on the protein (Tyr485CysfsTer27).

## DISCUSSION

β-mannosidosis is classified as a disorder of complex molecule degradation.^52^ Other disorders within this group include α-mannosidosis, fucosidosis, sialidoses, galactosialidoses, mucolipidoses, aspartylglucosaminuria, Kanzaki disease and Schindler disease.^30^ The gene *MANBA*, localized to chromosome 4q22-q25, comprises 136509 bp (NG_012804.2), 17 exons 34, and a transcript of 4001 bp (NM_005908.4) encoding an 879-amino acid protein ^36^ with a molecular weight of 100 kDa.^32^

Oligosacchariduria is a feature of β-mannosidosis^53^, and this finding is a consequence of the accumulation of partially degraded oligosaccharides derived from the degradation of N-glycans.^54^ This is the first publication describing the profile characteristic of this disorder by MALDI-TOF analysis, given that β-mannosidosis was not described in the initial publication of this method.^27^ Mannosyl-β(1 → 4)-N-acetylglucosamine and sialyl-α(2 → 6)-mannosyl-β(1 → 4)-N-acetylglucosamine have been previously described in urine of patients with β-mannosidosis ^1,12,55^, and both are present in the urine of Patients 1, 3 and 4. In Patient 2, only sialyl-α(2 → 6)-mannosyl-β(1 → 4)-N-acetylglucosamine was found, probably because of the effect of the transplant he received. In two patients (Patient 3 and Patient 4), a third abnormal peak was found, which corresponds to a Neu-Man2-GlcNac2 structure. This compound has not been described before in other affected patients, and it is present in the urine of the most severely affected patients in our cohort. One explanation is that these residues have not been degraded yet by chitobiase (endo-β-N-acetylglucosaminidase), an enzyme that removes the reducing N-acetylglucosamine, and leaves the oligosaccharide with only one terminal N-acetylglucosamine.^54^ The removal of the reducing N-acetylglucosamine is an essential step needed for the subsequent degradation of the N-glycan core by the specific α1–6 mannosidase, and the last degradation step by β-mannosidase.^56^ Chitobiase is present in rodents and primates, and absent in goats, which explains the more severe phenotype in these species.^57^ Additional studies are needed to delineate the relation between chitobiase and an abnormal β-mannosidase.

Establishing a genotype-phenotype correlation is difficult with the current knowledge. Except for the 13 patients from the Roma population, all the patients with reported variants have private mutations, and 9 patients (45%) are compound heterozygous. Two patients share a variant in the codon Arg182, which is located in the binding site of the protein. One of the patients is a compound heterozygous (Arg182Trp / Trp466Ter) female with angiokeratoma and mild hearing loss, without developmental delay or cognitive abnormalities.^31^ The newly published Patient 1 is a homozygous (Arg182Gln) male with hearing loss, learning difficulties, mild behavioral abnormalities, and normal development. Both patients who carry a missense variant in the position 182 of the protein have absent enzyme activity, and both have a milder phenotype. Riise Stensland et al.^40^ demonstrated that the Arg182Trp variant has no detectable enzyme activity in pCS2-transfected cells. This finding was also demonstrated with the Gly392Glu and Ser505Pro variants, which are associated with a mild^42^ and severe^39^ phenotypes, respectively; and they concluded that the mild phenotype is not explained by residual enzyme activity.^40^ Furthermore, the presence of residual activity also does not correlate with the severity of the disease manifestations. The patient described by He et al.^51^ was a compound heterozygous of a missense (p.Gln94Lys) and a frameshift (p.Met846IlefsTer27) variants, and residual enzyme activity in leukocytes was detected, although low for the reference range, but being the second highest residual activity detected from the cohort reported in literature (Table 1). This patient had a very severe phenotype given by recurrent pulmonary infections, cystic lesions, and alveolar hemorrhage, and died between 5-10 years of age due to fungal infection.

The variant c.2158-2A>G has been described in 13 patients, all of them from the Roma population.^14,18,49^ There is phenotypical variability in the affected patients, even in the same members of the family. From the four patients with reported enzyme activity, two had reduced enzyme activity, and in two the enzyme activity was completely absent (Table 1). Additionally, Kleijer et al. ^18^ described two affected siblings, one of them with coarse face, skeletal dysplasia, short stature, and severe and recurrent infections that caused her demise between 15-20 years of age. Her older sibling had intellectual disability, deafness, mild dysmorphic features, recurrent skin infections and normal stature, without skeletal dysplasia. Roma population is a highly endogamic ethnic group, and probably the variant c.2158-2A>G had a founder effect in this community. Safka Brozkova et al.^14^ calculated a heterozygous frequency of 3.77% in the Czech and Slovak normal hearing Roma population, but they did not find this variant in other Czech and Slovak non-Roma population evaluated through exome sequencing. Schrauen et al.^49^ found that Roma population from Hungary that carry the c.2158-2A>G variant have a divergent background between Asian and European, with a more pronounced South Asian ancestry, compared to other Roma groups which is similar to those individuals with European ancestry, therefore it is probable that the *MANBA* variant appeared in Roma population after they left India.^14^

In this study, we found that 43.3% of the patients have facial dysmorphism. Although facial coarsening can be seen in β-mannosidosis,^50^ this is not a common feature, and the dysmorphic facial features described in patients are usually mild. Also, dysostosis multiplex was seen only in 8% of the patients and organomegaly in 17.4%.

The neurological system is the most frequently affected in patients with β-mannosidosis, and this is manifested as the high incidence of intellectual disability (ID), behavioral abnormalities, and hearing loss. ID is a condition with a prevalence of 1-3% in the general population^58^ and its etiology is heterogeneous, although up to 40% of cases have a genetic cause.^59^ Up to 40% of patients with ID have a mental health problem, 39% of children with ID have ADHD, and 18% have autism.^60^ The proportion of patients with β-mannosidosis and behavioral abnormalities in this study is 84.6%, which suggests that the behavioral phenotype is caused by β-mannosidosis itself, and not secondary to ID. In three patients (6.8%) the acquisition of milestones was normal in the first year of age, and in the second year of life starts to slow down.^1,37,45^ Two patients have showed signs of regression of milestones: Patient 3 has developed urinary incontinence between 10 and 15 years old, and Patient 4 has severe dysphagia of new onset between 1-5 years old. There are no other reports of regression of milestones in other affected patients.

Twenty-five percent of the described patients had hearing loss as their first symptom (Table 2). From these patients, 95% have ID. On the other hand, 78.6% of the patients with intellectual disability have hearing loss (Fisher’s exact test p=0.156). There is one patient with hearing loss and angiokeratoma corporis diffusum without intellectual disability.^31^ In this scenario, we cannot rule out the possibility of underdiagnosis, especially in those cases of hearing loss with milder symptoms. β-mannosidosis should be considered as a differential diagnosis in those patients with syndromic and apparently non-syndromic hearing loss.

Table 3 shows other symptoms reported in patients with β-mannosidosis. As expected, behavioral abnormalities and neurological manifestations account for most of the reported symptoms. Interestingly, multiple eye manifestations have also been reported, including nystagmus in Patient 2. Yu et al.^61^ applied exome sequencing and bioinformatic methods to identify the genetic etiology for infantile nystagmus in Chinese population. The authors found the variants c.2013C>A, p.Arg638His and c.2346T>A, p.Leu749His in affected patients, both in heterozygous status. The patients did not show any other symptoms besides nystagmus, and the β-mannosidase enzyme activity was reduced between 40-70% in affected patients. The enzyme activity and the expression of *MANBA* in those two variants were reduced in transfected HEK293 cells. In the caprine model of β-mannosidosis, it has been demonstrated a decreased number of oligodendrocytes and a generalized myelin deficit in optic nerve that persists in 16-week-old animals, suggesting that the low amount of oligodendrocytes could be a consequence of abnormal stem cell proliferation or early death.^62^

**Table 3.**
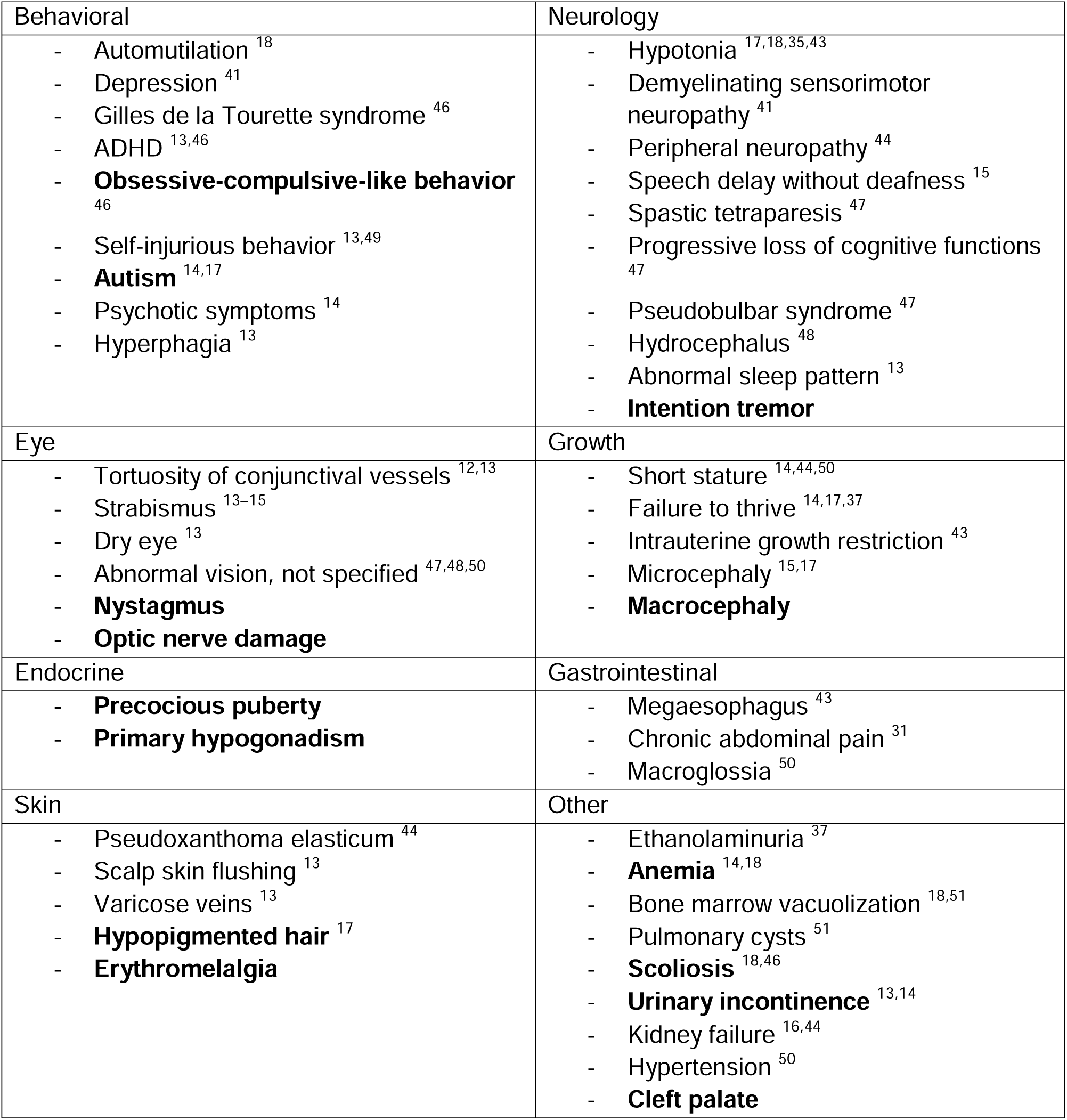
Other signs and symptoms previously reported and newly associated with β-mannosidosis. The signs and symptoms found in our patient cohort are in bold.

Another symptom not previously reported in β-mannosidosis is erythromelalgia in Patient 4. This condition is characterized by recurrent episodes of erythema, burning pain and increased temperature in distal extremities, and it has an incidence of 0.25-1.3 per 100.000 patients per year.^63^ Primary erythromelalgia is associated to pathogenic variants in *SCN9A* gene,^64^ which was ruled out in Patient 4. Secondary erythromelalgia has multiple etiologies, including polycythemia vera, essential thrombocytosis and autoimmune disorders.^63^ This condition has not been previously described in β-mannosidosis. The pathophysiology of erythromelalgia depends on its underlying etiology, and studies on this are scarce; however, the common finding is a central autonomic dysfunction secondary to abnormal sympathetic fibers, which causes impairment in the vasoconstrictor reflexes, along with small and large fiber neuropathy.^63,65^ A lysosomal storage disorder associated with chronic pain is Fabry disease, in which there is a progressive destruction of the nervous fibers due to accumulation of globotriaosylceramide in the lysosomes, causing cytoplasmic damage and cellular death.^66^ More studies should be developed to establish the pathophysiology of erythromelalgia in β-mannosidosis.

In this newest cohort, two different patterns of brain MRI imaging of β-mannosidosis were found. The first pattern, characterized by the delayed myelination in the first two years of Patient 4, and the lack of progression of myelination over time in Patient 2, is suggestive of hypomyelination^19,67,68^. The second pattern is visualized in the MRI of Patient 3, which shows hyperintensity on white matter in T1 with low T1 white matter signal. Interestingly, there is also cortical atrophy on the brain MRI in Patient 3. Hypomyelinating disorders are characterized by the deficit in myelin deposition, while dysmyelinating disorders are caused by the deposition of abnormal myelin.^67^ Other two types of oligosaccharidoses, fucosidosis and sialic acid disorders, also have hypomyelination as a manifestation on brain MRI, however, Barkovich and Deon^68^ consider it a secondary failure of myelination, and not a hypomyelinating disorder *per se* as seen in Pelizaeus-Merzbacher disease. Although β-mannosidosis is classified a genetic leukoencephalopathy ^69^, the pathophysiological mechanisms that explain the MRI findings are unknown in humans, and there is no literature about histopathological findings. The best studied animal models for β-mannosidosis are the caprine and bovine. In the caprine model, the main feature of brain damage is dysmyelination.^7^ Histologically, the neuronal tissue shows vacuolation of cells to various extents, as well as a reduction of oligodendrocytes and myelin paucity demonstrated by Luxol stain.^70^ The tissues that show the least amount of myelin are the corpus callosum, anterior commissure, rostral thalamic peduncle, hippocampus, cerebral peduncles, pyramids and brachium of the inferior colliculus, and these findings were consistent between the analyzed animals. Interestingly, in this study,^70^ the only cranial nerve that showed severe reduction of myelin was the eighth nerve. In another study,^71^ a complete absence of myelin sheaths was demonstrated in the corpus callosum, and in the areas with reduced amount of myelin, the number of oligodendrocytes was significantly reduced as well. The expression of proteolipid protein (PLP) is severely reduced in cerebral hemispheres (8% of normal) and brainstem (12% of normal), and the myelin fraction in brainstem and hemispheres is 2-7% of the normal in newborn and 4-week animals, demonstrating a severe deficit in myelination.^72^ Sasaki et al.^73^ showed that the myelin-associated glycoprotein (MAG) was also reduced, but its glycosylation pattern was not affected, however, the myelin basic protein (MBP) was not as severely affected as the other myelin proteins evaluated (MAG, PLP and CNP), and they hypothesized that there would be interference from the accumulated oligosaccharides to the transport system when myelin is synthesized.

Based on animal research and Renaud’s report, we hypothesize that the reason for the brain MRI findings is a low amount of myelination that does not change through time. Additional studies are needed to determine the pattern of myelination in β-mannosidosis.

## CONCLUSION

β-mannosidosis is a disorder that causes a neurological phenotype in association with heterogeneous manifestations, with a wide range of severity that cannot be explained only by a genotype-phenotype correlation. The low incidence of this condition may be due to underdiagnosis of those patients with milder phenotypes. We also provide evidence that brain imaging in β-mannosidosis can be manifested as hypomyelination or as abnormal white matter changes, associated with progressive cortical atrophy in severe cases. A new oligosaccharide structure was found in urine of affected patients. Oligosaccharide degradation plays an important role in the myelination of the central nervous system; therefore, studies are necessary to delineate the role of oligosaccharide metabolism in the pathophysiology of β-mannosidosis.

## DATA AVAILABILITY

The data that support the findings of this study is available upon request.

## ACKNOWLEDGEMENTS

The authors thank the patients and their parents for their participation in this study. AM thanks Sanofi-Genzyme for funding the Lysosomal Storage Diseases fellowship.

## AUTHOR CONTRIBUTIONS

Conceptualization: A.M., V.K.; Data curation: A.M., L.H.G., P.L.H.; Formal analysis: A.M.; Investigation: K.M.S., P.L.H., R.W., N.R.P., T.C.L., P.J.O., V.K.; Resources: K.M.S., K.H., P.L.H., G.B.P., R.W., N.R.P., P.O., V.K.; Visualization: A.M., L.H.G.; Writing-original draft: A.M.; Writing-review and editing: A.M., K.M.S., L.H.G., K.H., P.L.H., G.B.P., R.W., N.R.P.,T.C.L., P.J.O., V.K.

## ETHICS DECLARATION

This study was approved by the University of California Irvine Institutional Review Board (IRB #2024-3829). All patients’ parents or legally authorized representative provided written consent and provided us with their medical records, results of previous biochemical and genetic testing, and brain MRI images and results. The patients’ parents or legally authorized representative provided written consent for using photographs on this manuscript.

## SUPPLEMENTAL FIGURES

**Figure S1.** Missense MANBA variants found in the affected patients. For each variant, the upper image shows the alignment and the affected amino acid position in red square. The lower image shows the 3d structure (left: wild type protein, right: abnormal protein). **A:** Arg182Gln variant from Patient 1. **B.** Arg500His variant from Patient 2. **C**. Tyr477Cys and **D**. Gln145Pro variants from Patient 4.

**Figure S2.** Brain MRI of affected patients with β-mannosidosis. **A.** Patient 1 with normal brain MRI. From left to right, the projections are sagittal T1, axial T1, axial T2 and axial FLAIR. **B**. Patient 5, brain is normal. From left to right, brain coronal T1 and brain axial T2 projections. **C.** Patient 6 showing a normal brain MRI. From left to right, sagittal T1, axial T1, axial T2 and coronal T2 projections.

