## Supplemental table 1 for "Expanding the phenotype spectrum of β-mannosidosis"

**Table supl. 1**. Specific symptoms reported in each patient published. (+): symptom reported and present. (-): Symptom reported and absent. NR: symptom not reported. DD: Developmental delay. ID: Intellectual disability. HL: Hearing loss. AK: Angiokeratoma. RI: Recurrent infections. FD: Facial dysmorphism. SD: Skeletal dysplasia. BA: Behavioral abnormalities. OM: Organomegaly. SZ: Seizures. AT: Ataxia.

|  | **Author and year** | **Age of diagnosis** | **Age of symptoms onset** | **First symptom** | **DD** | **ID** | **HL** | **AK** | **RI** | **FD** | **SD** | **BA** | **OM** | **SZ** | **AT** |
| --- | --- | --- | --- | --- | --- | --- | --- | --- | --- | --- | --- | --- | --- | --- | --- |
| 1 | Cooper, 1986 | 40-45 years | 1-5 years | Intellectual disability | (-) | (+) | NR | (+) | NR | (-) | (-) | NR | (-) | NR | (-) |
| 2 |  | 15-20 years | NR | Intellectual disability | NR | (+) | NR | (+) | NR | NR | NR | NR | NR | NR | NR |
| 3 | Wenger, 1986 | 1-5 years | 1-5 years | Speech delay | (-) | (+) | (+) | NR | NR | (+) | (-) | NR | (-) | NR | (-) |
| 4 | Dorland, 1988 | 6-10 years | 0-1 year | Feeding difficulties and diarrhea | (-) | NR | (+) | (-) | (+) | (-) | (-) | (+) | (-) | NR | (-) |
| 5 |  | 6-10 years | NR | Feeding difficulties and recurrent infections | (-) | NR | (+) | (-) | (+) | (-) | (-) | (+) | (-) | NR | (-) |
| 6 | Kleijer, 1990 | N/A | NR ("early life") | Developmental delay | (+) | (+) | NR | NR | (+) | (+) | (+) | (+) | (-) | NR | NR |
| 7 |  | 26-30 years | NR | NR | NR | (+) | (+) | NR | (+) | (+) | (-) | (+) | NR | NR | NR |
| 8 | Wijburg, 1992 | 1-5 years | 1-5 years | Speech delay | (-) | (+) | (+) | NR | (+) | NR | NR | NR | (-) | (-) | (-) |
| 9 | Cooper, 1991 | 10-12 months | 0-1 year | Developmental delay | (+) | (+) | (-) | NR | NR | (-) | NR | NR | NR | (+) | (-) |
| 10 | Poenaru, 1992 | 1-5 years | NR | Speech delay | (-) | (+) | (-) | NR | (+) | (+) | (-) | (+) | (-) | NR | (-) |
| 11 | Levade, 1994 | 11-15 years | 6-10 years | Apathy, disinterest. Hand weakness (13 y) | (-) | (-) | (-) | (-) | (-) | (-) | (-) | (-) | (-) | NR | (-) |
| 12 | Rodriguez-Serna, 1996 | 20-25 years | 11-15 years | Angiokeratoma corporis diffusum | (-) | (-) | (-) | (+) | NR | (-) | (-) | (-) | (-) | (-) | (-) |
| 13 | Gourrier, 1996 | 7-9 months | 0-1 year | Chronic dysphagia | (+) | (+) | NR | (-) | (+) | (-) | (-) | NR | (+) | NR | (-) |
| 14 | Uchino, 2002; Suzuki, 2004 | 50-55 years | 1-5 years | Hearing loss | (+) | (+) | (+) | (+) | NR | (-) | (-) | NR | (-) | NR | (+) |
| 15 | Cherian, 2004 | 1-5 years | 0-1 year | Seizures | (+) | (+) | (-) | NR | NR | (-) | (-) | NR | (-) | (+) | NR |
| 16 |  | 1-5 years | 0-1 year | Seizures | (+) | (+) | (-) | NR | NR | (-) | (-) | NR | (-) | (+) | NR |
| 17 | Sedel, 2006 | 15-20 years | 1-5 years | Hearing loss | (-) | (+) | (+) | NR | (+) | (-) | (-) | (+) | (-) | (-) | (-) |
| 18 | Gort, 2006 | 20-25 years | NR | Angiokeratoma corporis diffusum | (-) | (-) | (+) | (+) | NR | NR | NR | (-) | NR | (-) | (-) |
| 19 | Molho-Pessach, 2007 | 36-40 years | Childhood | Intellectual disability | NR | (+) | (+) | (+) | (+) | NR | NR | (+) | NR | NR | NR |
| 20 | Labauge, 2009 | 15-20 years | 1-5 years | Intellectual disability | (-) | (+) | (-) | (-) | NR | (-) | (-) | NR | NR | NR | (+) |
| 21 | Broomfield, 2013 | 4-6 months | At birth | Seizures and hearing loss | (+) | (+) | (+) | NR | NR | (+) | NR | NR | (+) | (+) | (-) |
| 22 | Blomqvist, 2019 | 10-15 years | At birth | Hypotonia | (+) | (+) | (+) | (+) | (+) | (+) | NR | (+) | NR | NR | (-) |
| 23 | Schrauen, 2019 | NR | NR | Hearing loss | NR | (+) | (+) | NR | (+) | NR | NR | (+) | NR | NR | NR |
| 24 | Safka Brozkova, 2020 | NR | NR | NR | (+) | (+) | (+) | (-) | (+) | (-) | (-) | (+) | (-) | (-) | (-) |
| 25 |  | NR | NR | NR | (+) | (+) | (+) | NR | (-) | (+) | (-) | (+) | NR | NR | (-) |
| 26 |  | NR | NR | NR | NR | (+) | (+) | NR | NR | NR | NR | (+) | NR | NR | NR |
| 27 |  | NR | 1-5 years | Hearing loss | NR | (+) | (+) | NR | (+) | NR | NR | NR | NR | NR | NR |
| 28 |  | NR | NR | NR | (+) | (+) | (+) | NR | (+) | NR | NR | (+) | NR | NR | (-) |
| 29 |  | NR | NR | NR | NR | (+) | (+) | NR | (-) | (+) | NR | (+) | NR | NR | (-) |
| 30 |  | NR | NR | NR | NR | NR | (+) | NR | NR | NR | NR | NR | NR | NR | NR |
| 31 |  | NR | NR | NR | NR | (+) | (+) | NR | NR | NR | NR | (+) | NR | NR | NR |
| 32 |  | NR | NR | NR | NR | NR | (+) | NR | NR | NR | NR | NR | NR | NR | NR |
| 33 |  | NR | NR | NR | NR | NR | (+) | NR | NR | NR | NR | NR | NR | NR | NR |
| 34 |  | NR | NR | NR | NR | NR | (+) | NR | NR | NR | NR | NR | NR | NR | NR |
| 35 | Alshoraim, 2021 | 1-5 years | NR | NR | (+) | (+) | (+) | NR | NR | (+) | (+) | (+) | NR | NR | NR |
| 36 | Gowda, 2021 | 1-5 years | Late infancy | Developmental delay | (+) | NR | (-) | NR | NR | (-) | (-) | (+) | (+) | NR | (-) |
| 37 |  | 1-5 years | NR | Developmental delay | (+) | NR | (-) | NR | NR | (-) | (-) | (+) | (-) | NR | (-) |
| 38 | He, 2022 | 1-5 years | Infancy | Recurrent infections | (+) | NR | NR | NR | (+) | (-) | (-) | (-) | (-) | NR | (-) |
| 39 | This publication | 6-10 years | 1-5 years | Hearing loss | (-) | (+) | (+) | (-) | (-) | (-) | (-) | (+) | (-) | (-) | (-) |
| 40 |  | 1-5 years | 0-1 years | Developmental delay | (+) | (+) | (-) | (-) | (-) | (+) | (-) | (+) | (-) | (-) | (+) |
| 41 |  | 6-10 years | 1-5 years | Frequent infections | (-) | (+) | (+) | (-) | (+) | (+) | (-) | (+) | (-) | (+) | (-) |
| 42 |  | 1-5 years | Newborn | Failed hearing newborn screening | (+) | (+) | (+) | (+) | (+) | (+) | (-) | (+) | (-) | (+) | (-) |
| 43 |  | 10-15 years | 1-5 years | Hearing loss | (-) | (+) | (+) | (+) | (-) | (+) | (-) | (+) | (+) | (-) | (-) |
| 44 |  | 26-30 years | 1-5 years | Hearing loss | (+) | (+) | (+) | (+) | (-) | (-) | (-) | (-) | (-) | (-) | (+) |
|  | **Mean (SD)** | 12.8 years (±13.4) | 28 months (±34.1) | **Symptoms present (%)** | 18 (56.3) | 32 (91.4) | 29 (74.4) | 10 (52.6) | 17 (70.8) | 13 (41.9) | 2 (7.4) | 23 (82.1) | 4 (16) | 6 (40) | 4 (13.8) |
|  |  |  |  | **Symptoms absent (%)** | 14 (43.8) | 3 (8.6) | 10 (25.6) | 9 (47.4) | 7 (29.2) | 18 (58.1) | 25 (92.6) | 5 (17.9) | 21 (84) | 9 (60) | 25 (86.2) |
|  |  |  |  | **Total reported patients** | 32 (72.7) | 35 (79.5) | 39 (88.6) | 19 (43.2) | 24 (54.5) | 31 (70.5) | 27 (61.4) | 28 (63.6) | 25 (56.8) | 15 (34.1) | 29 (65.9) |
|  |  |  |  | **Not reported (%)** | 12 (27.3) | 9 (20.5) | 5 (11.4) | 25 (56.8) | 20 (45.5) | 13 (29.5) | 17 (38.6) | 16 (36.4) | 19 (43.2) | 29 (65.9) | 15 (34.1) |
